# Utilization of Pharmacy Value-Added Services and Its Association with Waiting Time for Medication Collection in Public Health Institutions across Malaysia

**DOI:** 10.1101/2022.02.28.22271668

**Authors:** Mazuin Mahmud Taridi, Muhammad Radzi Abu Hassan, Rozita Mohamad, Chan Huan Keat, Noraini Nordin, Chan Pui Lim, Ong Chin Wen, Khairul Iman Muzakir, Azuana Ramli

## Abstract

**Background:** Pharmacy value-added services (PVAS) have long been offered in public health institutions across Malaysia as an alternative to conventional counter services for prescription refills, with the aim to reduce the waiting time.

**Objective:** To assess the utilization of the PVAS in individual health institutions, and its association with the achievement of the key performance indicator (KPI) set for the pharmacy waiting time.

**Method:** This was a cross-sectional study based on the data contributed by 142 hospitals and 648 health clinics throughout 2018. The availability and uptake of the PVAS were summarized as percentages. The impacts of the PVAS uptake and the other institution-related factors on the KPI achievement were further explored using the logistic regression analysis.

**Results:** Approximately 2.9 million (17.1%) of the refill prescriptions were dispensed via the PVAS. The appointment-and-pickup services (42.7%) and the Integrated Drug Dispensing System (23.7%) emerged as the most commonly used types of PVAS. A higher PVAS uptake was associated with a better KPI achievement (OR=0.91, 95% CI: 0.84-0.98). In contrast, adding a new type of PVAS to the existing services yielded an opposite outcome (OR=1.48, 95% CI: 1.15-1.89). Both the prescription load and location of health institutions were also found have influenced the KPI achievement.

**Conclusion:** The PVAS are generally well accepted in Malaysia and showed to have reduced the pharmacy waiting time. However, strategies to optimize the PVAS uptake are warranted.

## INTRODUCTION

Universal access to high-quality healthcare is the ultimate goal of public health systems worldwide. One of the strategies to improve the quality of healthcare is by finding a way to reduce the waiting time for health services^1^. Prolonged waiting time in health institution is always associated with poor patient satisfaction, which is also believed to have increased the stress level and affected the health-related quality of life, particularly among the chronically ill population^2^. Therefore, a wide range of innovative measures, including the use of an automated prioritization system, have been taken not only to ease the congestion in health institutions but also to prioritize the care for vulnerable populations^3^.

In the context of pharmacy practice, waiting time typically refers to the time required for patients to collect their medications from the pharmacy counter following a medical consultation, or to get a refill of their prescriptions^4^. Apart from reflecting poorly on the efficiency of healthcare providers, prolonged waiting time in a pharmacy setting could adversely affected their commitment to their treatment plan^5^. While providing sufficient information upon dispensing is crucial to ensure medication adherence in patients,^6^ it remains a challenge for pharmacists to professionally perform such a task under time pressure. In addition to causing a long waiting time at the pharmacy department, the conventional counter dispensing services might have also inconvenienced some patients by requiring them to make additional trips to health institutions for refills^7,8^Other than that, longer waiting time at pharmacy counter could possibly due to manual prescription by doctors which make it challenging for the pharmacist to read as well as at some facilities the pharmacy’s counter and space are too small as compared to the number of patients they have to serve^9^.

As the pharmacy services are likely to have a great impact on the overall impression of the health system, the waiting time at the outpatient pharmacy department has long been made by the Ministry of Health (MOH) as one of the key performance indicators (KPIs) for all the public hospitals and clinics in Malaysia as well as the Client Charter in MoH facilities. It is mandated that at least 95% of prescriptions, both new and refill, be filled and dispensed within 30 minutes at the outpatient pharmacy department^10^. However, the increasing patient load constantly stands in the way of meeting the standard set for the KPI. In 2017 alone, the public health institutions in Malaysia received a total of 58.7 million prescriptions, approximately 12.2% higher than the number of prescriptions received in the previous year^11^.

In the wake of such limitations, the MOH has been impelling for the use of a series of novel services, collectively known as the Pharmacy Value-Added Services (PVAS), for the prescription refills since the last decade^11,12^. The common types of PVAS made widely available in public hospitals and clinics across the country include (i) the appointment-and-pickup services, which allow patients to set an appointment date for their subsequent refills by using either an appointment card or various modes of communication, such as phone calls, short message service (SMS), communication applications, fax and email, (ii) the Integrated Drug Dispensing System (IDDS), a nationwide standard referral system allowing patients to get their refills from a preferred health institution near to the patients house under the Ministry of Health, Malaysia, (iii) the drive-through services, which allow patients to pick up their refills without having to park or leave their vehicles, and (iv) the medication delivery by post services, which allow the refills directly sent to where a patient lives or work at minimal postal charges^12^. Meantime, a number of health institutions have also been making efforts to initiate new forms of PVAS to complement the conventional pharmacy counter services.

As compared with the similar services provided in other countries,^14,13^ the strength of the PVAS lies in the variety of the services offered, each of which could be tailored according to the capacity of an institution and the need of a local community. As much as the PVAS are expected to improve the patient satisfaction, the uptake of the services is voluntary and patients are still allowed to opt for conventional counter services to pick up their monthly refills. Although the last few years witnessed an increase in the number of refill prescriptions dispensed via the PVAS,^15^ the association between such an initiative and the pharmacy waiting time for medication collection remains unclear. The existing studies on the PVAS are generally limited by the their single-center design and a relatively small sample size^12,16^. As efforts and resources have been put in by public health institutions in Malaysia to initiate the PVAS, this study was designed to assess the utilisation of the services and its association with the achievement of the KPI set for the pharmacy waiting time on a nationwide scale.

## METHODS

### Study Design and Data Source

This was a retrospective cross-sectional study on the utilization of the PVAS and how it influenced the waiting time for medication collection in 2018. The findings were generated based on the data collected by the Pharmaceutical Services Program (PSP) from the public health institutions, both hospitals and health clinics, which were known to have provided at least one type of the PVAS in 2018. The health institutions which were not staffed with pharmacists or pharmacy assistants, and those managing only acute and uncomplicated cases which did not require patients to refill their prescriptions, were excluded. The implementation of the PVAS had been centrally coordinated and monitored by the PSP since they were launched. Each institution offering the PVAS was required to submit report on the utilization of the services quarterly. Meanwhile, the PSP has also been closely monitoring the waiting time at the outpatient pharmacy departments of all the public health institutions. In this context, the pharmacy waiting time referred to the duration between the point at which a prescription was received at a pharmacy counter and the point at which patients were called to pick up their medications^17^.

### Data Collection and Assessment

This study was registered and approved with the National Medical Research Register (NMRR) Malaysia (NMRR-19-3040-50122). The data was directly obtained from the PSP dataset. A data collection form was constructed to gather the following information of each health institution: (i) the prescription load (the number of new prescriptions, refill prescriptions and items per prescription), (ii) the capacity (the number of counters and pharmacy staff), (iii) utilization of the PVAS (the types of PVAS offered and the uptake of each service), (iv) the average waiting time for medication collection and the achievement of the KPI, and (v) the setting (hospital or clinic) and location (Northern Peninsula, Central Peninsula, Southern Peninsula, East Coast or East Malaysia).

The PVAS uptake in each health institution was calculated in relation to the total number of refill prescriptions received in the same year. According to their prescription load, the health institutions were grouped into group 1 hospitals, group 2 hospitals, group 1 health clinics and group 2 health clinics. Half of the hospitals and health clinics, which had the higher prescription load in 2018 (median), were grouped under the group 1 hospitals and health clinics, while the lower were grouped as group 2 respectively.

### Statistical Analysis

The statistical analysis was performed using the SPSS for Windows version 21.0 (IBM, New York). All the categorical variables were summarized as frequencies and percentages, and numerical variables as means and standard deviations (SDs), or as medians and interquartile ranges (IQRs), as appropriate. The association between the PVAS uptake and the failure to achieve the KPI set for the pharmacy waiting time, along with the roles of other institution-related factors, was further explored by using the simple and multiple logistic regression (backward stepwise method) analyses. The results were presented as odds ratios (ORs), along with their corresponding 95% confidence intervals (CIs) and p-values. The final model was also tested for multicollinearity and interactions, while its fitness was confirmed through the Hosmer-Lemes how goodness-of-fit test, the overall correctly classified percentage and the area under the receiver operating characteristic curve. The significant level of the statistical test was set at 5%.

## RESULTS

A total of 790 public health institutions in Malaysia, including 142 hospitals and 648 health clinics, were found to have provided at least one type of PVAS in 2018. They were widely distributed across different regions of the country (98 in the Northern Peninsula, 190 in the Central Peninsula, 187 in the Southern Peninsula, 189 in the East Coast, and 126 in East Malaysia). Over the one-year period, all the public health institutions had received a total of 55,407,903 prescriptions. Hospitals contributed to nearly 40% of the overall prescription load, more than 80% of which fell on the group 1 hospitals. Similarly, the prescriptions from the group 1 health clinics composed approximately 80% of the overall prescription load of health clinics. The proportion of refill prescriptions ranged from 23.8% to 33.2% across different settings (Figure 1).

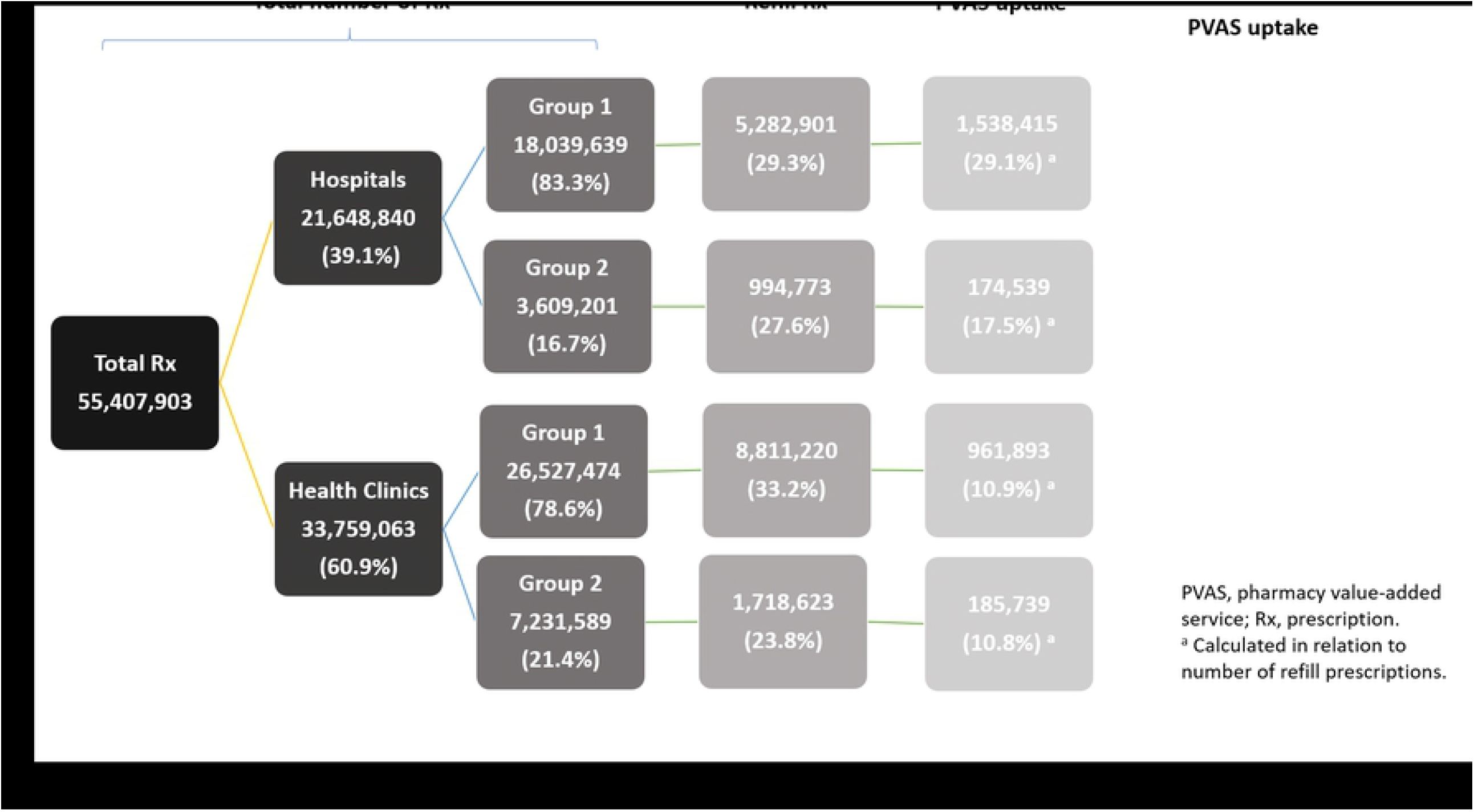

On average, the health institution was staffed with four pharmacists and pharmacy assistants, and had two dispensing counters. More than 90% of them provided the appointment-and-pickup services, and slightly more than 80% implemented the IDDS. Apart from the four major types of the PVAS, more than one-third of them also provided alternative services, such as the medication locker and home medication review services. Most group 1 hospitals provided at least three types of PVAS, while other settings mainly provided only one or two types of services (Table 1).

**Table 1:** Capacity, availability of PVAS and waiting time of outpatient pharmacy departments at four major groups of public health institutions across Malaysia, 2018.

| Variables | Overall<br>(n=790) | Hospitals |  | Health clinics |  |
| --- | --- | --- | --- | --- | --- |
|  |  | Group 1<br>(n=71) | Group 2<br>(n=71) | Group 1<br>(n=324) | Group 2<br>(n=324) |
| **Number of pharmacy staff <sup>a</sup>, median (IQR)** | 4.00 (5.00) | 14.00 (13.00) | 5.00 (3.00) | 6.00 (4.00) | 3.00 (2.00) |
| **Number of dispensing counters, median (IQR)** | 2.00 (1.00) | 4.00 (2.00) | 2.00 (1.00) | 2.00 (1.00) | 1.00 (0.00) |
| **Types of PVAS offered, n (%)** |  |  |  |  |  |
| IDDS | 639 (80.9) | 71 (100.0) | 70 (98.6) | 301 (92.9) | 197 (60.8) |
| Appointment-and-pickup services | 729 (92.3) | 70 (98.6) | 65 (91.5) | 306 (94.4) | 288 (88.9) |
| Drive-through services | 57 (7.2) | 24 (33.8) | 2 (2.8) | 29 (9.0) | 2 (0.6) |
| Medication delivery by post services | 162 (20.5) | 53 (74.6) | 12 (16.9) | 89 (27.5) | 8 (2.5) |
| Miscellaneous | 298 (37.8) | 59 (83.1) | 29 (40.8) | 168 (51.9) | 42 (13.0) |
| **Number of PVAS types offered, n (%)** |  |  |  |  |  |
| 1 | 169 (21.4) | 0 (0.0) | 3 (4.03) | 21 (6.5) | 145 (44.8) |
| 2 | 322 (40.8) | 6 (8.5) | 40 (56.3) | 128 (39.5) | 148 (45.7) |
| 3 | 169 (21.4) | 21 (29.6) | 18 (25.4) | 102 (31.5) | 28 (8.6) |
| 4 | 90 (11.4) | 22 (31.0) | 9 (12.7) | 56 (17.3) | 3 (0.9) |
| 5 | 35 (4.4) | 18 (25.4) | 1 (1.4) | 16 (4.9) | 0 (0.0) |
| 6 | 5 (0.6) | 4 (5.6) | 0 (0.0) | 1 (0.3) | 0 (0.0) |
| **Waiting time for medication collection, minutes, mean (SD)** | 10.95 (5.63) | 15.85 (4.58) | 12.81 (4.53) | 13.43 (5.19) | 6.99 (3.69) |
| **Meeting the standard of the KPI for waiting time, n (%)** | 635 (80.4) | 51 (71.8) | 62 (87.3) | 208 (64.2) | 314 (96.9) |

IQR, interquartile range; IDDS, Integrated Drug Dispensing System; KPI, key performance indicator; PVAS, pharmacy value-added service; SD, standard deviation.
<sup>a</sup> Including pharmacists and pharmacy assistants.

The overall PVAS uptake was 17.1%. The group 1 hospitals recorded the highest PVAS uptake (29.1%), followed by the group 2 hospitals (17.5%) and the group 1 health clinics (10.9%) (Figure 1). The most widely used PVAS type was the appointment-and-pickup services, followed by the IDDS and the drive-through services. Such a trend cut across different settings, except for the group 1 hospitals, in which the IDDS were the most frequently used services (Figure 2).

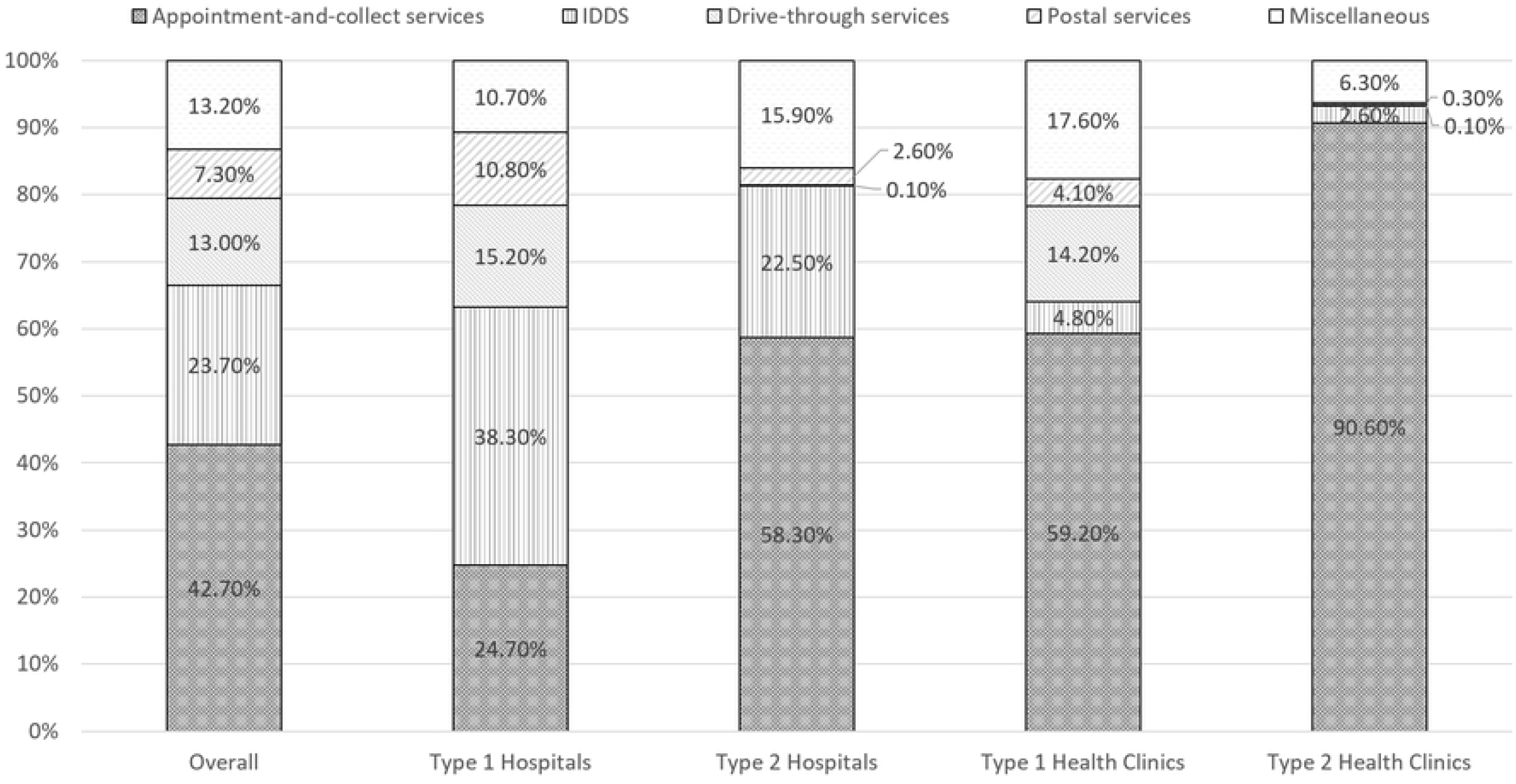

The health institutions recorded an average pharmacy waiting time of 10.95 minutes for medication collection, and 80.4% of them met the standard set for the KPI (Table 1). As compared with the group 2 health clinics, the group 1 hospitals (adjusted OR: 4.25; 95% CI: 1.51, 11.98) and the group 1 health clinics (adjusted OR: 10.90; 95% CI: 5.26, 22.56) were shown to have a higher tendency of not achieving the KPI goal. A higher PVAS uptake increased the likelihood of hitting the target (adjusted OR: 0.91; 95% CI: 0.84, 0.98). However, offering more types of services resulted in an opposite outcome (adjusted OR: 1.48; 95% CI: 1.15, 1.89) (Table 2). In comparison with the health institutions located in the Northern Peninsula, those located in East Malaysia (adjusted OR: 2.64; 95% CI: 1.33, 5.22) were more likely to fail in meeting the standard of the KPI. In contrast, health institutions in the East Coast had a lower risk of not achieving the KPI goal (adjusted OR: 0.39; 95% CI: 0.18, 0.84).

**Table 2:** Factors associated with the failure to achieve the standard set for the key performance indicator of the pharmacy waiting time, logistic regression analysis.

| Variables | Simple logistic regression |  |  | Multiple logistic regression <sup>a</sup> |  |  |
| --- | --- | --- | --- | --- | --- | --- |
|  | b | OR (95% CI) | p | b | OR (95% CI) | p |
| **Setting** |  |  |  |  |  |  |
| Group 1 hospital | 2.51 | 12.31 (5.45, 27.81) | <0.001 | 1.45 | 4.25 (1.51, 11.98) | 0.006 |
| Group 2 hospital | 1.52 | 4.56 (1.78, 11.68) | 0.002 | 0.90 | 2.45 (0.89, 6.75) | 0.084 |
| Group 1 health clinic | 2.86 | 17.51 (8.97, 34.20) | <0.001 | 2.39 | 10.90 (5.26, 22.56) | <0.001 |
| Group 2 health clinic | 0 | 1 | - | 0 | 1 | - |
| **Number of PVAS types offered** |  |  |  |  |  |  |
|  | 0.08 | 1.76 (1.51, 2.01) | <0.001 | 0.39 | 1.48 (1.15, 1.89) | 0.002 |
| **Percentage of refill prescriptions dispensed using PVAS** |  |  |  |  |  |  |
|  | -1.37 | 0.99 (0.94, 1.04) | 0.659 | -0.10 | 0.91 (0.84, 0.98) | 0.012 |
| **Location <sup>b</sup>** |  |  |  |  |  |  |
| Northern Peninsula | 0 | 1 |  | 0 | 1 |  |
| Central Peninsula | -0.13 | 0.88 (0.50, 1.55) | 0.658 | -0.13 | 0.88 (0.47, 1.64) | 0.685 |
| Southern Peninsula | -0.54 | 0.58 (0.32, 1.05) | 0.073 | -0.31 | 0.73 (0.38, 1.43) | 0.363 |
| East Coast | -1.45 | 0.23 (0.12, 0.48) | <0.001 | -0.93 | 0.39 (0.18, 0.84) | 0.016 |
| East Malaysia | 0.34 | 1.41 (0.78, 2.54) | 0.253 | 0.97 | 2.64 (1.33, 5.22) | 0.005 |

b, regression coefficient; OR, odds ratio; CI, confidence interval.
<sup>a</sup> Multicollinearity and interaction term were checked and not found. Hosmer-Lemeshow test (p=0.201), classification table (overall correctly classified percentage=80.8%) and area under curve (82.4%) were applied to check the model fitness.
<sup>b</sup> Health institutions were grouped according to their locations (states) into Northern Peninsula (Perlis, Kedah, Penang), Central Peninsula (Perak, Selangor and Kuala Lumpur), Southern Peninsula (Negeri Sembilan, Melaka and Johor), East Coast (Kelantan, Terengganu and Pahang), and East Malaysia (Sabah, Sarawak and Labuan).

## DISCUSSION

This study clearly indicates that the PVAS, a set of novel pharmacy services introduced by the MOH in Malaysia, are getting increasingly well accepted among the patients. The number of prescription dispensed by the PVAS was shown to have reached a historic high at 2,860,586 (17.1%) in 2018, higher than that reported in 2014 (14%)^13^. To the best of our knowledge, this is the first situational analysis of the PVAS on a nationwide scale. Aside from the utilization of the PVAS, the findings also provide insight into how the services could affect the waiting time, which has been an important quality indicator of the pharmacy services as a whole.

The appointment-and-pickup services emerged as the most widely available and frequently used services. Such findings imply that this particular type of PVAS has been highly feasible and sustainable in resource-limited settings. It is arguably the least cost-consuming PVAS type and easy to set up, as it only requires a telephone or any other communication devices, which enable patients to make an appointment and collect their refills upon their arrival. It has also been generally well accepted by patients due to its potential to significantly reduce the waiting time, as demonstrated by Othman et al. The relatively high uptake of the appointment-and-pickup services also suggests that patients still highly value the opportunities to meet with the pharmacy staff and receive advices on medications in person, as much as they desire not to wait long in front of the pharmacy counters.

Instead of the appointment-and-pickup services, the IDDS was shown to be the most widely used PVAS type in the group 1 hospitals. Such findings would be expected as most group 1 hospitals typically serve as referral centers for a wide range of specialties, and are not necessarily the most preferred settings for patients to pick up their refills. Additionally, the group 1 hospitals were also found to have an exceptionally high prescription load, approximately five times higher than that of their group 2 counterparts. This also explains why group 1 hospitals recorded the longest pharmacy waiting time among all the settings. Therefore, it is conceivable that the group 1 hospitals would highly recommend the uptake of the IDDS to patients. Patients are also likely to opt for the IDDS, simply because they do not need to regularly return to high-volume hospitals. By addressing all these limitations, the IDDS allows patients to pick up their refills from settings, which are less congested and nearer to their residence or workplace.

However, the drive-through and medication delivery by post services, which were initially designed to address the car parking problems in health institutions, were not widely used as they would be expected. The drive-through services have the potential to ease the congestion at the pharmacy counters and benefit the vulnerable populations.^18^ However, the feasibility of such services is primarily limited by the need for extra costs and space to build the facilities, as well as additional manpower to run the service. Therefore, the drive-through services are not currently made available for medication collection in many health institutions. On the other hand, the relatively low uptake of the medication delivery by post services is likely attributable to the postal fees, which could be deemed as a burden, particularly by patients of a lower socioeconomic status. It is also likely that patients are not well informed about the services, mainly due to inadequate promotion from providers^17^. Additionally, the existing post services in Malaysia also do not allow the delivery of psychotropics and photo- and heat-sensitive medications, and are not available in some remote areas due to limited postage coverage by the logistic partner courier company.

Despite their limitations, this study also shows that increasing the PVAS uptake had increased the achievement of the KPI set for the waiting time. A similar finding was reported by Loh et al ^19^ where the report concluded that the patient waiting time in the ambulatory pharmacy is improved as the VAS registration increased. Nevertheless, such findings should not be mistaken for a reason to vary the services in a health institution, as the logistic regression model also indicates that adding new types of PVAS to what are already available could, conversely, lengthen the waiting time.

Although it is understandable that some health institutions have been taking the initiative to diversify their services to meet the need of different populations, the findings of this study suggest that they should be more strategic in the selection of services to provide, particularly by taking their own capacity into the consideration without compromising the KPI achievement. This is reasonable, as in resource-limited public hospitals and health clinics, initiating a new service is likely to augmented the manpower shortage and consequently affect the operation of the conventional pharmacy counter services. Thus, instead of introducing new types of PVAS, emphasis should be placed on optimizing the uptake of existing PVAS in the future. In our record, at least one-third of the health institutions were found to have offered unique and unclassifiable PVAS other than those highly recommended by the MOH, however further investigation into the effectiveness of such a strategy is thus required.

Even though the standard set for the KPI was fulfilled by slightly more than 80% of the health institutions, it is noteworthy that approximately 30% of the group 1 hospitals and 35% of the group 1 health clinics still did not achieve the targeted waiting time for medication collection. As compared with the group 2 health clinics, the group 1 health clinics even demonstrated approximately 11 times higher odds of not achieving the KPI set for the pharmacy waiting time. Given that approximately 7 in 10 prescriptions were still dispensed through the conventional counter services in the high-volume health institutions in Malaysia, efforts to upscale the promotion of the PVAS use are necessary^11^. At the same time, the reasons for geographical differences in the waiting time for medication collection remain questionable, calling for further research in this area.

The strength of this study lies in the representativeness of its findings, as the data was obtained directly from the PSP, which has long been centrally monitoring the implementation of the services across the country. As positive as the findings of this study sound, the roles of many factors which could also have an impact on the achievement of the KPI, such as the resources put into the services, the efficiency of individual health institutions, the medication inventory management and the extent to which the electronic hospital information system was used to facilitate the medication dispensing, were not comprehensively explored. Furthermore, it was assumed that all the data used in this study was collected and reported according to the guidelines set by the PSP by individual health institutions.

## CONCLUSION

After a constant initiative for patient-centered services for years, it is encouraging to note the uptake of the PVAS for medication collection, which were launched by the MOH more than a decade ago, had reached 17.1% in 2018 and 22.3% in 2019. Increasing the PVAS uptake was shown to have led to a better achievement of the KPI set for the waiting time in a public health institution. Hence, continuous efforts to scale up the use of PVAS, especially in a busy hospitals and health clinics throughout the country, are warranted. Going forward, besides the waiting time, the potential roles of the PVAS in reducing the medication wastage and improving the patient compliance and satisfaction, as well as their economic implications, could also be explored.

## Data Availability

Data cannot be shared publicly because this is the government dataset. Data are available from the Malaysia Ministry of Health Data Access for researchers who meet the criteria for access to confidential data

## Acknowledgements and Funding

This research received no specific grant from any funding agency in the public, commercial, or not-for-profit sectors. We would like to thank the Director General of Health Malaysia for his permission to publish this article. We would like to acknowledge Dr. Abdul Haniff bin Mohamad Yahaya dan Ms. Siti Fauziah binti Abu for their support and suggestions to improve this manuscript.

